# Evaluation of the Implementation of the Safewards Model in an Open-Door Acute Inpatient Unit. A quasi-experimental study

**DOI:** 10.64898/2025.12.03.25340056

**Authors:** Anna Moreno-Orea, Laura Miró Mezquita, Joan de Pablo Rabasso, Tatiana Bustos Cardona, Antonio R. Moreno-Poyato, Zaida Agüera, Maria Teresa Lluch-Canut, Jorge Cuevas-Esteban

**Author notes:** Corresponding author**: Antonio R. Moreno-Poyato.** Departament d’Infermeria de Salut Pública, Salut Mental i Maternoinfantil, Facultat d’Infermeria, Universitat de Barcelona. Calle Feixa Llarga s/n, 08907, L’Hospitalet de Llobregat, Barcelona, Spain., **Zaida Agüera**. Departament d’Infermeria de Salut Pública, Salut Mental i Maternoinfantil, Facultat d’Infermeria, Universitat de Barcelona. Calle Feixa Llarga s/n, 08907, L’Hospitalet de Llobregat, Barcelona, Spain.

## Abstract

**Background:** Safewards is a multi-intervention mental health nursing model evidence-based practice, aimed at preventing and reducing conflict and containment.

**Objective:** To evaluate the effectiveness of the implementation of Safewards model in an open-door acute mental health unit. This analysis examines the impact of the Safewards model on conflict levels and restrictive therapeutic measures.

**Design:** A prospective quasi-experimental study was conducted between August 2019 and January 2022, with patients admitted to an acute mental health unit from Spain.

**Methods:** Descriptive and inferential analysis will be conducted. Multivariate regression models will be used to investigate the relationship between coercive measures, conflict levels, and clinical and demographic variables, adjusting for covariates if necessary.

**Results:** The results indicated statistically significant differences in several variables such as rule violations (F=6.11, df= 1, p < .014), verbal aggression (F=14.9, df = 1, p < .001), and physical aggression towards objects (F = 7.2, df= 1, p < .008).

The results indicated a statistically significant differences in restrictive therapeutic measures: intramuscular forced medication (F=8.5, df= 1, p < .004), environmental seclusion (F=14.6, df = 1, p < .001), and verbal/behavioural containment (F=75.0, df = 1, p < .001).

**Conclusion:** This is the first study to evaluate the implementation of the Safewards model in an open-door acute inpatient unit in Spain with respect to restrictive therapeutic measures.

**Social media abstract:** Safewards Model in an Open-Door Acute Inpatient Unit in Barcelona @annabcn_24, @SafewardsBCN, @jcuevasesteban

**What is already known?:**

- Safewards reduces conflict and restrictive interventions in mental health inpatient units.
- Safewards uses 10 interventions based on mental health nursing theory.
- Safewards is a mental health model for practice improvement implemented internationally.

**What this paper adds:**

- This study is the first evaluation of Safewards’ implementation in Spain.
- The research team translated Safewards into Spanish for staff training.
- Translating Safewards aids its use in Spanish-speaking mental health units.
- Restrictive measures reduced significantly, except for mechanical restraint.

**Reporting Method:** This study adheres to the relevant EQUATOR guidelines and follows the STROBE (Strengthening the Reporting of Observational Studies in Epidemiology) reporting method.

**Patient or Public Contribution:** The implementation of the Safewards model relies on the active participation of both healthcare professionals and patients to promote a safer and more therapeutic environment. Through structured interventions, staff facilitate communication strategies, de-escalation techniques, and conflict resolution, while patients actively contribute to fostering a climate of mutual respect and support. This bidirectional approach not only reduces coercion and incidents but also strengthens the therapeutic relationship and enhances the sense of community within the unit

## INTRODUCTION

The Safewards model was developed with the aim of reducing conflicts and the use of coercive therapeutic measures in acute mental health inpatient units (Bowers et al., 2015; Dickens et al., 2020). Safewards is a care model comprising 10 interventions grounded in mental health nursing theory (Bowers, 2014). The model focuses on modifying factors that influence the relationship between healthcare staff and patients to reduce the likelihood of conflictual events (Bowers et al., 2014).

Within the context of the model, “conflict” refers to user behaviors that threaten their security or the security of others, including aggressive conduct, suicide attempts, self-harm, substance use, and absconding (Baker et al., 2009). It also encompasses non-compliance with basic rules, such as refusing therapy or smoking in prohibited areas, which may generate tensions with staff or other patients (Bowers et al., 2005). On the other hand, the term “containment” in Safewards is used to describe the therapeutic strategies employed by staff to manage difficulties in the unit’s dynamics. These strategies include forced medication (oral or intramuscular), increased supervision or observation and seclusion (Bowers et al., 2007).

The model identifies six areas of potential factors that can trigger critical situations in acute mental health inpatient units. These factors arise from six “Domains of Origin”: patient characteristics, regulatory framework, staff team, physical environment, external elements outside the hospital, and the patient community. Each of these domains can generate specific points of tension for both staff and patients, thus increasing the likelihood of conflicts and restrictive measures. Safewards also suggests a dynamic and reciprocal relationship between conflict and containment.

The model proposes that staff in inpatient units can influence conflict and containment rates at four levels:

1. By reducing or eliminating the factors that give rise to conflicts, thereby preventing points of tension from emerging.
2. By disrupting the link between points of tension and conflict, ensuring that the latter does not translate into conflictual events.
3. By consciously choosing not to use containment in situations where it may be counterproductive.
4. By ensuring that the use of containment does not generate further conflict when it is applied (Kingston et al., 2011).

Since Safewards primarily focuses on what staff can do, the model suggests that early intervention in the “Domains of Origin” can prevent problems before they escalate into critical points (Hahn et al., 2013).

## BACKGROUND

Research studies on the implementation of the Safewards care model have been conducted in various countries such as the United Kingdom (Bowers et al., 2014; Davies et al., 2020; James et al., 2017), Germany (Baumgardt et al., 2019), Denmark (Stensgaard et al., 2018), Sweden (Björkdahl et al., 2013), Australia (Fletcher et al., 2017), Canada (Kipping et al., 2019; Whitmore, 2017), Poland (Lickiewicz et al., 2021), Finland (Asikainen et al., 2020), and the United States (Leveillee et al., 2019). The Safewards model has undergone extensive evaluation across various mental health settings, including adult (Fletcher et al., 2017) adolescent (Fletcher et al., 2017), geriatric (Dawson et al., 2024) forensic (Maguire et al., 2022; Price et al., 2016) and intellectual disability services (Kipping et al., 2019; Riding, 2016). Recent pilot studies have extended Safewards implementation to emergency department settings (Gerdtz et al., 2021). Evidence has reported reductions ranging from 2% to 40% in the use of coercive measures (Baumgardt et al., 2019; Björkdahl et al., 2013; Bowers, 2014; Bowers et al., 2015; Fletcher et al., 2017; Stensgaard et al., 2018). However, the effectiveness of Safewards is contingent upon several factors, including the prior experience of healthcare staff, their acceptance of the model, and their willingness to implement it (Björkdahl et al., 2013; Fletcher et al., 2017; James et al., 2017).

Currently, there is an ongoing debate about whether closed-door units may increase the incidence of violent incidents and absconding. Longitudinal prospective studies conducted in countries such as Switzerland (Hochstrasser et al., 2018; Jungfer et al., 2014) and Germany (Huber et al., 2016) have demonstrated that open-door units reduce the number of absconding incidents, self-harm behaviours, and restrictive measures such as seclusion, mechanical restraint (Krückl et al., 2023), and pharmacological interventions, including forced medication. A recent systematic review published in 2020 (Gooding et al., 2020) indicates that the Safewards model and open-door policies in inpatient settings, alongside the Six Core Strategies, have shown the greatest effectiveness in preventing and reducing conflictual events, as well as the use of restrictive measures.

Scientific evidence (Efkemann et al., 2019; Fletcher, Hamilton, et al., 2019; Whittington et al., 2023) supports the implementation and evaluation of care models centred on therapeutic relationships and person-centred care in open acute mental health units, as well as models aimed at reducing conflict and the consequent use of coercive measures. To our knowledge, no evaluation of the implementation of the Safewards model has been conducted in acute psychiatric inpatient units within the Spanish healthcare context, nor in open-door units at an international level. Therefore, the main objective of this study was to assess the effectiveness of the Safewards model in reducing conflict and containment in an acute mental health hospitalization unit.

## METHODS

### Study design

A prospective quasi-experimental study was conducted between August 2019 and January 2022.

### Setting and participants

The Safewards model was implemented at the psychiatry acute unit of <<anonymized information>>.

The psychiatry department consists of an emergency area and an inpatient unit which with 14 beds for acute inpatient admission. The unit reported an occupancy rate of 89.72% in 2019 and 87.13% in 2021. During the baseline period, the total number of discharges was 72, followed by 60 discharges during the implementation phase, and 58 discharges during the consolidation phase. The study involved the participation of nurses, auxiliary nursing care technicians, healthcare assistants, psychiatrists, clinical psychologists, and a social worker.

The ward operates under open-door policies, characterized by the door remaining open during daytime hours to facilitate a more therapeutic and less restrictive environment. At night time, for safety and operational reasons, the door is closed. This approach balances accessibility and safety while fostering a sense of freedom and autonomy for patients during the day. These open-door policies were formally implemented starting in October 2021, reflecting a shift towards modern, patient-centered care practices designed to reduce the perceived institutional nature of inpatient mental health settings. This policy aligns with broader evidence suggesting that open-door practices can decrease incidents of absconding and reduce the need for restrictive measures, while promoting a culture of trust and collaboration within the unit.

### Data collection procedure

Data collection was carried out using the Patient–staff Conflict Checklist Shift Report (PCC-SR; Bowers *et al*., 2005). Data were collected via a form completed at the end of each work shift—morning, afternoon, and night.

The initial phase of data collection took place during the pre-implementation period from August to October 2019 to establish the baseline status of the unit. During this period, the research team translated the Safewards model into Spanish following a collaborative agreement with King’s College London and the Maudsley Hospital, with coordination from the creators of the care model (http://www.safewards.net/es/). In October 2019, a 15-hour training and standardisation course was delivered to the healthcare team, including nursing and medical professionals, followed by the subsequent implementation with the assignment of activity leaders.

In the second phase, the Safewards model was progressively implemented over 13 weeks, from November 2019 to January 2020.

The data collection and implementation timeline were affected by the COVID-19 pandemic, which led to the suspension of all psychiatric services at the hospital. The impact of the pandemic necessitated significant efforts to resume the project. The period from February to July 2021 was therefore excluded from the analysis.

Post-implementation data were analysed during a consolidation and sustainability phase, covering the period from November 2021 to January 2022, during which the implemented activities were maintained.

Ethical approval was obtained from the Research Ethics Committee (REC) of <<anonymized information>> (REC Reference: PI-22-231).

### Intervention

The 10 interventions of the Safewards model were implemented, including: clarifying mutual expectations, kind words, verbal de-escalation, positive words, getting to know each other, discharge messages, reassurance, mutual help meeting, relaxation techniques, and support following bad news. All staff received the same training, delivered across three workshops of five hours each, and the same support for implementing the activities during the initial preparation phase, the 13-week implementation phase (where activities were introduced progressively), and the consolidation phase.

Key factors considered during implementation included hospital management approval and operational task definition. and an analysis of the methodologies used in other studies. To facilitate this, an expert panel was established, comprising the head of the department, the nursing supervisor, the head of the acute unit’s medical section, and the clinical psychologist. A leader was assigned for each of the interventions, with responsibilities for planning, supervising, and supporting the implementation process.

A Safewards implementation committee was formed, consisting of the intervention leaders and the expert panel, serving as a platform to address challenges, discuss strategies, and propose improvements. The introduction of activities was staggered; during the first week, four activities were implemented, and by the end of the fourth week, seven had been introduced with a high level of adherence. The implementation of each intervention was monitored through weekly committee meetings.

Specific instructions were provided for the interventions, such as integrating them into daily routines and identifying the necessary materials for their execution, with some adaptations made to meet the unit’s specific needs. Clarifying mutual expectations and relaxation techniques presented the most difficulties during implementation. Monitoring of the degree of implementation was carried out using the “researcher visit fidelity checklist”(James et al., 2017), a tool used in the original Safewards trial, which employed a combination of self-reporting and ward visits by the research team. Complete descriptions of the interventions are freely available on the official website (www.safewards.net/es/).

### Measures

Two instruments were used for the assessment:

– Patient Staff Conflict Checklist (PCC-SR; (Bowers et al., 2005)), Spanish version. The Patient-Staff Conflict Checklist was employed to document the frequency of patient conflict behaviors (e.g. self-harm, absconding, violence, medication refusal) either attempted or successful, and the staff containment measures used to maintain safety (e.g. intermittent special observation, constant special observation, seclusion, physical restraint etc.) and was compiled using strict definitions at the end of every nursing shift. This is a tool developed by Bowers et al. (2005) and used routinely during the current investigation. Staff were trained in the use of the PCC-SR and provided with a handbook to refer to for definitions of different types of events. It was adapted to our clinical setting characteristics, including variables such as mechanical restraint, which is not used in the original version from United Kingdom. The incidents classified as “conflicts” measured during the study were grouped into subcategories, with analyses conducted for both the baseline and outcomes periods. These subcategories included incidents related to medication, rule violations, tobacco use, alcohol or substance use, attempted or successful escapes, verbal aggression, physical aggression towards objects, physical aggression towards people, suicide attempts or self-harm, and the total sum of incidents. All incidents associated with restrictive therapeutic measures, referred to in the Safewards model as “containment,” applied within the 13 weeks before and after the implementation period of the Safewards Model were analysed. Coercive interventions (Pollmächer, 2015) were defined as any actions taken against a patient’s will that limit their personal freedom or harm their physical integrity. These were similarly grouped into subcategories: forced oral medication, forced intramuscular medication, environmental seclusion, verbal containment, mechanical restraint, and the total sum of interventions.
– Fidelity Checklist (Bowers et al., 2005). The Fidelity Checklist is a brief standardised audit tool used in the original Safewards trial (Bowers et al., 2015) in the United Kingdom to determine the degree of adherence to the interventions comprising the model. In the present study, the checklist designed by the team from Victoria, Australia (Fletcher et al., 2020) was used, which assesses the implementation of each activity following an observation of the inpatient unit by members of the evaluation team. During this process, observations and inquiries were made with the healthcare team to complete the checklist. Approximately 30 minutes were spent completing the quantitative and qualitative elements of the Fidelity Checklist. The checklist was scored out of 10 to reflect the number of interventions being implemented and to analyse the percentage of implementation for each activity. The principal investigator completed the fidelity checklist. Weekly monitoring occurred during the first 18 weeks of implementation, followed by monthly monitoring.

### Statistical analysis

SPSS v.29 for Windows was used for the statistical analysis. The comparison between the groups was performed using the Chi-square test for categorical variables, and analysis of variance (t-student) for quantitative variables. The correction for Type-I error due to multiple statistical significance tests was performed using Bonferroni’s method.

## RESULTS

The primary outcomes of the study were counts of conflict and containment obtained through the PCC recording. A total of 404 PCC forms were completed by nursing staff at the end of each work shift. The response rate was 74% of PCC forms filled out. There were fewer missing data in the baseline phase (11.4%; 242/273) than in the outcome phase (40.7%; 162/273; χ^2^ = 60.912, df = 1, p < 0.001). The outcome was the count of conflict and containment events per shift in the inpatient unit (morning from 8 am to 3 pm, afternoon from 3 pm to 10 pm, and night from 10 pm to 8 am). There were significantly fewer missing outcome data from night shifts (156/182; 14.3% missing) compared with morning (136/182; 25.3% missing) and afternoon (112/182; 38.5% missing) shifts (overall analysis, χ^2^ = 27.715, df = 2, p < 0,001).

The Chi-square test revealed significant associations between the completion rates of the PCC logs in each shift (χ² = 60.912, p < .001), indicating that the distribution of records during the baseline and outcome periods differed significantly across categories.

Subsequently, a student’s *t*-test was conducted to compare differences in conflicts and subcategories among several groups. The results indicated statistically significant differences in several variables such as rule violations (F=6.11, df=1, p < .014), verbal aggression (F=14.9, df = 1, p < .001), and physical aggression towards objects (F = 7.2, df= 1, p < .008).

Similarly, a student’s *t*-test was conducted to assess differences in restrictive therapeutic measures and subcategories among several groups. The results indicated a statistically significant differences in the variables: intramuscular forced medication (F=8.5, df=1, p < .004), environmental seclusion (F=14.6, df = 1, p < .001), and verbal/behavioural containment (F=75.0, df = 1, p < .001).

### Conflict and contention rates throughout implementation

In the comparison of the incidence of “conflict”, significant differences were observed across the various categories. Results showed that verbal aggression and physical aggression towards objects were significantly reduced, while non-compliance with rules increases. For the remaining conflict events, no statistically significant differences were found (see Table 1).

**Table 1.** Incidence of conflict by subcategories.

| Incidents | Baseline<br>n=242 | Post-implementation<br>n=162 | Total<br>n=404 | Statistics |  |  |
| --- | --- | --- | --- | --- | --- | --- |
|  |  |  |  | F | df | p |
| Medication incidents, mean (SD) | n=427<br>1.76<br>(1.56) | n=262<br>1.62 (1.61) | n=689<br>1.71<br>(1.58) | 0.843 | 1 | .36 |
| Noncompliance with rules, mean (SD) | n=258<br>1.07<br>(1.73) | n=279<br>1.72 (3.54) | n=537<br>1.33<br>(2.63) | 6.112 | 1 | .014 |
| Alcohol or substance use, mean (SD) | n=3<br>0.01<br>(0.11) | n=0<br>0.00 (0.00) | n=3<br>0.01<br>(0.09) | 2.023 | 1 | .16 |
| Escape attempts or escapes, mean (SD) | n=1<br>0.00<br>(0.06) | n=0<br>0.00 (0.00) | n=1<br>0.00<br>(0.05) | 0.669 | 1 | .41 |
| Verbal aggression, mean (SD) | n=96<br>0.40<br>(1.26) | n=2<br>0.01 (0.11) | n=98<br>0.24<br>(1.00) | 14.933 | 1 | <.001 |
| Physical aggression against objects, mean (SD) | n=33<br>0.14<br>(0.65) | n=0<br>0.00 (0.00) | n=33<br>0.08<br>(0.50) | 7.218 | 1 | .008 |
| Physical aggression against people, mean (SD) | n=11<br>0.05<br>(0.31) | n=1<br>0.01 (0.08) | n=12<br>0.03<br>(0.24) | 2.562 | 1 | .11 |
| Suicide attempts or self-harm, mean (SD) | n=2<br>0.01<br>(0.09) | n=0<br>0.00 (0.00) | n=2<br>0.00<br>(0.07) | 1.343 | 1 | .25 |
| Total incidents | n=831<br>3.43<br>(3.35) | n=544<br>3.36 (4.41) | n=1375<br>3.40<br>(3.81) | 0.038 | 1 | .85 |

*Note: This table presents the mean and standard deviation (SD) for each type of incident during the baseline and outcome phases, along with statistical tests (F, degrees of freedom (df), and p-values) assessing the differences between phases.*

In terms of the incidence of restrictive therapeutic measures, a significant reduction in all interventions was observed except for mechanical restraint. During the outcome period, no events were recorded for restrictive therapeutic measures related to forced oral or intramuscular medication (see Table 2).

**Table 2.** Incidence of Restrictive Therapeutic Measures.

*Incidence of Restrictive Therapeutic Measures*
| Interventions | Baseline<br>n=242 | Post-implementation<br>n=162 | Total<br>n=404 | Statistics |  |  |
| --- | --- | --- | --- | --- | --- | --- |
|  |  |  |  | F | df | p |
| Oral forced medication, mean (SD) | n=25<br>0.10<br>(0.61) | n=0<br>0.00 (0.00) | n=25<br>0.06<br>(0.48) | 4.604 | 1 | .032 |
| Intramuscular forced medication, mean (SD) | n=14<br>0.06<br>(0.25) | n=0<br>0.00 (0.00) | n=14<br>0.03<br>(0.20) | 8.595 | 1 | .004 |
| Environmental seclusion, mean (SD) | n=25<br>0.10<br>(0.34) | n=0<br>0.00 (0.00) | n=25<br>0.06<br>(0.27) | 14.650 | 1 | <.001 |
| Verbal containment, mean (SD) | n=468<br>1.93<br>(2.68) | n=15<br>0.09 (0.47) | n=483<br>1.20<br>(2.28) | 75.037 | 1 | <.001 |
| Mechanical restraint, mean (SD) | n=2<br>0.01<br>(0.09) | n=5<br>0.03 (0.39) | n=7<br>0.02<br>(0.26) | 0.743 | 1 | .39 |
| Total interventions | n=534<br>2.21<br>(2.87) | n=20<br>0.12 (0.61) | n=554<br>1.37<br>(2.48) | 82.610 | 1 | <.001 |

*Note: This table presents the mean and standard deviation (SD) for each type of incident during the baseline and outcome phases, along with statistical tests (F, degrees of freedom (df), and p-values) assessing the differences between phases.*

### Control of fidelity

The activities were gradually implemented with weekly fidelity monitoring for each activity. The overall fidelity was 67%.

At the audit conducted at the end of the implementation phase, the overall fidelity to the model was 72.50%. The level of adherence increased in almost all activities by the end of the consolidation period, where a fidelity of 80.50% was achieved.

**Figure 1:**
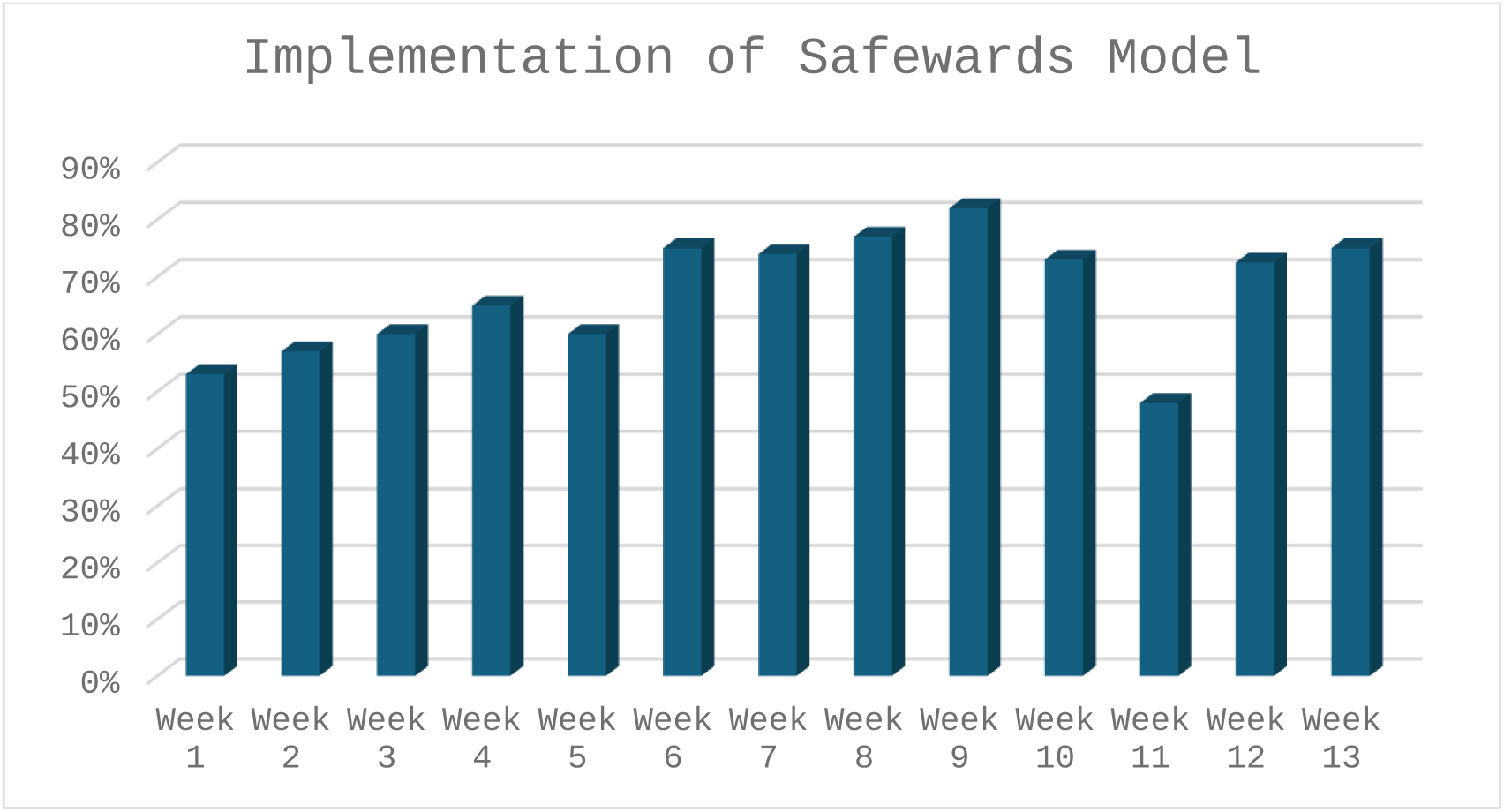
Fidelity of the implementation of Safewards.

## DISCUSSION

To the best of our knowledge, this is the first study that aims the implementation effectiveness of the Safewards care model in an open-door acute inpatient unit for reducing conflicts and restrictive therapeutic measures. The main findings of the present study suggest that implementing Safewards in the acute unit reduces conflicts and restrictive therapeutic measures.

In the present study, the model was not implemented as in the original trial, which had an initial period of 8 weeks; instead, a 13-week period was used, which was necessary for the implementation of the activities and defined the duration of the different phases. In a study conducted in two closed units in Germany (Baumgardt et al., 2019), the frequency and duration of coercive interventions applied during an 11-week period before and 11 weeks after the implementation period were analysed, and fidelity to the Safewards model was assessed using the Organisation Fidelity Checklist.

Extensive observations and repeated consecutive measurements of restrictive therapeutic measures were obtained before and after applying the Safewards model. These results derive from a prospective analysis of historical data collected in a natural setting rather than a controlled research environment, offering a realistic perspective on the intervention’s practical effects The PCC, developed by Bowers (Bowers et al., 2015) in the randomised clinical trial in the UK, allowed for the evaluation of Safewards’ impact on various significant indicators of conflict and containment. In our study, the PCC completion rate was 74%, being significantly higher during the night shift, followed by the morning shift, and lastly, the afternoon shift. These completion rates were greater than those reported in other previous studies, such as the original Safewards study from UK, where a 53% record completion rate was indicated (Bowers et al., 2015) or a study conducted in Australia, where a registration rate of 63.2% was obtained (Dickens et al., 2020). Our high registration rates suggest that automating record-keeping routinely at the end of each shift, following the guidelines of the Price et al. study (2016), is beneficial for compliance performance. In a prior study conducted in Australia (Fletcher, Hamilton, et al., 2019), the use of the PCC was deemed unfeasible, as it was considered too time-consuming for staff to complete. The good results achieved in our study may be related to the weekly meetings held with representatives from all work shifts, addressing difficulties in record-keeping, resolving doubts, and providing daily supervision until full completion was achieved.

Regarding conflict reductions, our findings showed a significant reduction in conflict rates during the implementation of the Safewards model, particularly in verbal aggression and physical aggression towards objects. These results are similar to those found in previous studies conducted in Australia (Maguire et al., 2018), where fewer conflict incidents were reported after the model’s implementation. As in the present study, all conflicts were grouped into a variable called “total conflict incidents”. They are also in line with a study performed in Canada (Dickens et al., 2020), where the authors found that the overall conflict rate was reduced by 23.0% compared to initial the baseline data after the implementation of Safewards. Overall, our findings reinforce previous results indicating the effectiveness of this model for the reduction of conflict episodes in psychiatric admission units.

Training in verbal de-escalation, one of Safewards’ activities, resulted in a significant reduction in related incidents (Brenig et al., 2023).

Regarding the frequency of restrictive therapeutic measures, our findings also suggest a significant reduction in nearly all of them compared to the weeks prior to Safewards’ implementation, except for mechanical restraint. Several studies (Fletcher *et al*., 2017; Stensgaard *et al*., 2018; Kipping, De Souza and Marshall, 2019; Dickens, Tabvuma and Frost, 2020; Lickiewicz *et al*., 2021 Bowers *et al*., 2015), also demonstrated a lower incidence of violence and coercive measures following the implementation of Safewards, while others showed no significant effect (Ward-Stockham et al., 2022). Our findings are consistent with previous studies (Baumgardt et al., 2019; Bowers et al., 2015; Dickens et al., 2020; Fletcher et al., 2017; Johnson, 2010) that also found a reduction in these measures. We found an overall reduction of 30.77% in coercive measures. Although our rates of reduction are lower than those reported in a previous study in Sweden (Pelto-Piri et al., 2024) hat indicated levels of reduction in coercive measures of up to 75%, our data are similar to those reported in other studies that found reduction rates of 24% in the UK (Bowers et al., 2015) and the 36% reported in studies in Australia (Maguire et al., 2018). However, our results are not in line with other studies (Stensgaard et al., 2018) in which the authors did not find changes in restraint episodes after the implementation. The contradictory findings could be explained by the low acceptance and adherence of staff to the intervention, unlike in ours. Additionally, the positive changes obtained in our study could also be linked to routine follow-ups with established meetings, supervision, and adequate training, as described in the Baumgardt study (Baumgardt et al., 2019). Moreover, our study benefits from the strength of having baseline measures for comparison. As Johnson et al. (2010) pointed out, most studies on the use of restraints have not employed comparison groups alongside experimental ones. In this study, we followed an evaluation process recommended by Bowers, which involved comparing data with those from a comparable prior period.

We found that the rate of forced medication, both oral and intramuscular, showed a significant decrease. This finding is consistent with a retrospective study conducted in Denmark (Stensgaard et al., 2018).

Another relevant finding is the lack of reduction in the use of mechanical restraints. Our result differs from studies conducted in Poland (Lickiewicz et al., 2021) and Australia (Baumgardt et al., 2019) which reported a decrease in both the use and duration of mechanical restraints following the implementation of Safewards. These results are supported by factors facilitating the implementation of Safewards, such as low staff turnover, professional experience, resilience, and optimal material resources (Baumgardt et al., 2019; Wilson et al., 2018). Our findings align with those of other studies that did not observe a reduction in the use of mechanical restraints (Maguire et al., 2018; Stensgaard et al., 2018).

A possible explanation for the lack of significant effect on the mechanical restraint rate since Safewards’ implementation may lie in the existing political context and trends. Human rights have emerged as a central focus in evaluating mental health policies and strategies (Herrman & Swartz, 2007; Patel et al., 2007), and the World Health Organisation (WHO) urges all nations to safeguard the rights of people hospitalised with mental disorders (Scobie et al., 2006) and the use of mechanical restraint in psychiatric units. Given that mechanical restraint rates were already low before Safewards’ implementation, achieving further reductions might be challenging.

Finally, regarding fidelity, the implementation of the Safewards Model in our centre demonstrated a high degree of adherence, consistent with other studies that also observed high fidelity (Baumgardt et al., 2019; Bowers et al., 2015; Fletcher et al., 2017). However, it is important to note that fidelity assessment is based solely on objective and visible evidence of the model’s application. This evaluation does not account for staff commitment to the model’s principles, their attitude towards it, or the difficulties encountered during implementation (Higgins et al., 2018). Therefore, fidelity results do not reflect whether staff have internalised the concepts of the Safewards Model or to what extent. Furthermore, they do not provide information on the model’s effectiveness in everyday clinical practice. These aspects should be addressed in future studies.

In research published in Australia, patients described feelings of hope, safety, respectful relationships, a sense of community, and reported feeling “more connected” with care staff after implementing the Safewards care model (Fletcher, Buchanan-Hagen, et al., 2019; Maguire et al., 2018). Other researchers also highlight increased nurse-patient interaction as a strategy to reduce conflict events (Lantta et al., 2016), enhancing a sense of safety and improving the ability to distract from difficult thoughts and feelings (Pelto-Piri et al., 2024).

### Limitations

Our study should be considered in the framework of several limitations. First, a major limitation is that the study design did not include a control group. It would have been optimal to include a control group to conduct a separate analysis for the intervention and control groups, allowing for a comparison of changes in the use of coercive measures. Additionally, the results were affected by the COVID-19 pandemic. There was a pause in the model’s implementation within the inpatient unit, and months were excluded from the analysis until the unit regained its functionality. Another significant limitation is that the sample size was too small to permit detailed analysis. Nevertheless, the methodology used for data collection adds rigour to our findings and contributes to the growing evidence base for interventions in acute mental illnesses within international health services. Patient participation is a crucial component of implementing Safewards, preferably as part of an ongoing co-creation process between staff and patients (Clifton et al., 2013; Kipping et al., 2019; Voorberg et al., 2015). Since many Safewards interventions require active patient involvement, the responsiveness of patients is a critical factor for implementation, which should be addressed in further studies.

Despite these limitations, the study also presents notable strengths. Firstly, to the best of our knowledge, this is the first study conducted in an open-door acute psychiatric inpatient unit of a major public hospital in Spain. The results are based on a prospective analysis of data generated in a natural setting outside a controlled research environment. This provides a realistic view of the expected impact of the interventions’ implementation in clinical practice, offering empirical evidence on the effectiveness of the Safewards programme within the Spanish healthcare system. Secondly, the translation of the Safewards model into Spanish facilitates staff training in Spanish-speaking countries and its implementation in mental health units.

## Conclusions

The current study supports previous publications asserting that the Safewards model is an effective intervention used in acute mental health inpatient units, significantly reducing conflicts and restraint measures. Adherence to the model and a higher degree of fidelity to its activities play a crucial role in achieving these outcomes. The model implementation fosters changes in how healthcare professionals interact with patients, marking a shift from a distant approach to a more engaged one.

Routine conflict and containment measurements per shift offer valuable data for intervention evaluation and should be widely adopted. A randomized controlled trial would be a relevant design for future studies on the effect of implementing the Safewards model in psychiatric inpatient units.

Future research could also involve various inpatient units and different types of patients to further evaluate the generalizability and effectiveness of the Safewards model across different settings.

## Relevance to Clinical Practice

Conflicts of interest: all authors declare no conflict of interest

## Supporting information

Ethics committee

## Data Availability

All data produced in the present study are available upon reasonable request to the authors

## Acknowledgments

We extend our gratitude to the care team involved in the implementation of the model, and especially to the nursing team for their dedicated hours, commitment, and valuable contributions, which have been an invaluable aid to our study.

