## Supplementary material for "Evaluation of the Implementation of the Safewards Model in an Open-Door Acute Inpatient Unit. A quasi-experimental study": Ethics committee

Germans Trias i Pujol
Hospital

Comitè d'Ètica de la Investigació

Institut Català
de la Salut

Ethics committee of Hospital Universitari Germans Trias i Pujol gave ethical approval for this work.

REF.CEI PI-22-231

2022/10/07

Anna Moreno Orea

Data: 2025.12.03 10:46:51 +01'00'
